# No Relationship between Male Pubertal Timing and Depression – New Insights from Epidemiology and Mendelian Randomization

**DOI:** 10.1101/2023.05.22.23290322

**Authors:** Raphael Hirtz, Corinna Grasemann, Heike Hölling, Nicola Albers, Anke Hinney, Johannes Hebebrand, Triinu Peters

## Abstract

**Background:** In males, the relationship between pubertal timing and depression is understudied and less consistent than in females, likely for reasons of unmeasured confounding. To clarify this relationship, a combined epidemiological and genetic approach was chosen to exploit the methodological advantages of both approaches.

**Methods:** Data from 2,026 males from a nationwide study (KiGGS) were used to investigate the non-/linear relationship between pubertal timing defined by the age at voice break and major depressive disorder (MDD), considering a multitude of potential confounders and their interactions with pubertal timing. This analysis was complemented by Mendelian Randomization (MR), which is robust to inferential problems inherent to epidemiological studies. We used 71 single nucleotide polymorphisms related to pubertal timing in males as instrumental variable to clarify its causal relationship with MDD based on data from 807,553 individuals (246,363 cases and 561,190 controls) by univariable and multivariable MR, including BMI as pleiotropic phenotype.

**Results:** Univariable MR indicated a causal effect of pubertal timing on MDD risk (inverse-variance weighted: OR=0.93, 95%-CI [0.87–0.99)], p=.03). However, this was not confirmed by multivariable MR (inverse-variance weighted: OR=0.95, 95%-CI [0.88–1.02)], p=.13), consistent with the epidemiological approach (OR=1.01, 95%-CI [0.81–1.26], p=.93). Instead, the multivariable MR study indicated a causal relationship of BMI with MDD by two of three methods.

**Conclusions:** Pubertal timing is not related to MDD risk in males. Considering the adverse health outcomes of higher BMI levels, our findings support the rationale for preventive measures to address obesity and its related risk factors.

## Introduction

Major depressive disorder (MDD) is a debilitating psychiatric disorder and is expected to be the leading cause of disease burden worldwide in 2030 by the WHO (Doherty, Egan, & Dinneen, 2021). Childhood and adolescent psychopathology, including MDD, relates to poor mental health and social functioning in later life (Fergusson, Boden, & Horwood, 2007), implicating the need to identify children and adolescents at risk for early prevention and intervention (Jones et al., 2018). While the prevalence of MDD rises during puberty and is higher in adolescent girls than boys (Mojtabai, Olfson, & Han, 2016), also the 12-month prevalence of MDD in adolescent boys in western countries ranges between 5.4% (Lu, 2019) and 5.7% (Mojtabai et al., 2016) and cumulatively amounts to 13.6% between 12 and 17 years of age (Breslau et al., 2017).

However, the relationship between puberty and MDD is less consistent in boys than in girls (Hamlat, Stange, Abramson, & Alloy, 2014; Wang, Lin, Leung, & Schooling, 2016), likely for reasons of unmeasured confounding (Wang et al., 2016). Recently, Stumper and Alloy (2021) reviewed a large number of potential confounders in this relationship, including BMI, body image, stressful and adverse life events, and parental and social support. However, some studies did not account for these confounders (Angold, Costello, & Worthman, 1998; Hamlat, McCormick, Young, & Hankin, 2020), and many only included a (small) subset.

To consider the methodological limitations of epidemiological studies, especially the latter of residual confounding, the present study addresses the relationship between male pubertal timing (MPT) and MDD by Mendelian randomization (MR). MR uses genetic markers to draw causal conclusions on the association between an exposure (e.g., pubertal timing) and an outcome (e.g., MDD) of interest by exploiting that genotypes are not generally associated with confounders in the population and are randomly assigned at conception, analogous to randomization in clinical trials (Davies, Holmes, & Davey Smith, 2018). Moreover, since the individual genotype is determined upon conception and cannot be modified by the outcome of interest, MR is robust to reverse causation. Usually, single nucleotide polymorphisms (SNPs) derived from large-scale genome-wide association studies (GWASs) are used as instrumental variable (IV) to study the relationship between exposure and outcome.

However, also the MR approach comes with limitations. Two-sample MR studies that rely on summary statistics from independent GWAS on the exposure and the outcome (Burgess & Thompson, 2021) are often limited by studying the relationship of interest based on the statistical, usually linear model used in the GWAS to define the IV. Thus, it is also not possible to study interactions between the exposure and other phenotypes in multivariable MR. Moreover, MR studies are not suitable for estimating effect sizes for specific developmental episodes, as significant findings represent the average effect of an exposure on the outcome over the life course (Burgess & Thompson, 2021). Thus, this approach is complemented by a large-scale study of adolescents and young adults (‘The German Health Interview and Examination Survey for Children and Adolescents’ - KiGGS), also including information on the confounders mentioned above in the relationship between pubertal timing and MDD.

Using this combined methodological approach, the results of the present study may have implications at the population level to guide prevention measures that address the timing of puberty in boys. At the individual level, MPT may be considered part of a prospective risk score to determine the likelihood of MDD if identified as an important contributing factor.

## Epidemiological Analysis

### Participants

KiGGS is a national longitudinal study on the health status of children and young people in Germany. Since the baseline study (2003-2006, N = 17,640), two follow-ups have been completed: KiGGS wave 1 between 2009 and 2012 (N = 11,992; age range: 0-17 years) and KiGGS wave 2 between 2014 and 2017 (N = 10,853; 3,775 respondents did not take part in any follow-up assessment; age range: 10-31 years). Further details on the study design, sampling strategy, and study protocol have been described in detail elsewhere (Mauz et al., 2020).

The KiGGS study was approved by the ethics committee of the Charité Berlin (baseline and wave 1) and the ethics committee of the Hannover Medical School (wave 2). Written informed consent was obtained from parents as well as from children aged 14 years and older. For the present study, data from the baseline study and wave 2 were used. Male participants who were queried on MDD history were considered for analysis (N = 2,732). Participants with incomplete information on MDD history, a history of MDD before 9 years of age (i.e., before pubertal onset is expected in boys), precocious or delayed puberty, and missing information on pubertal development or any confounder of interest were excluded (Figure 1).

**Figure 1.**
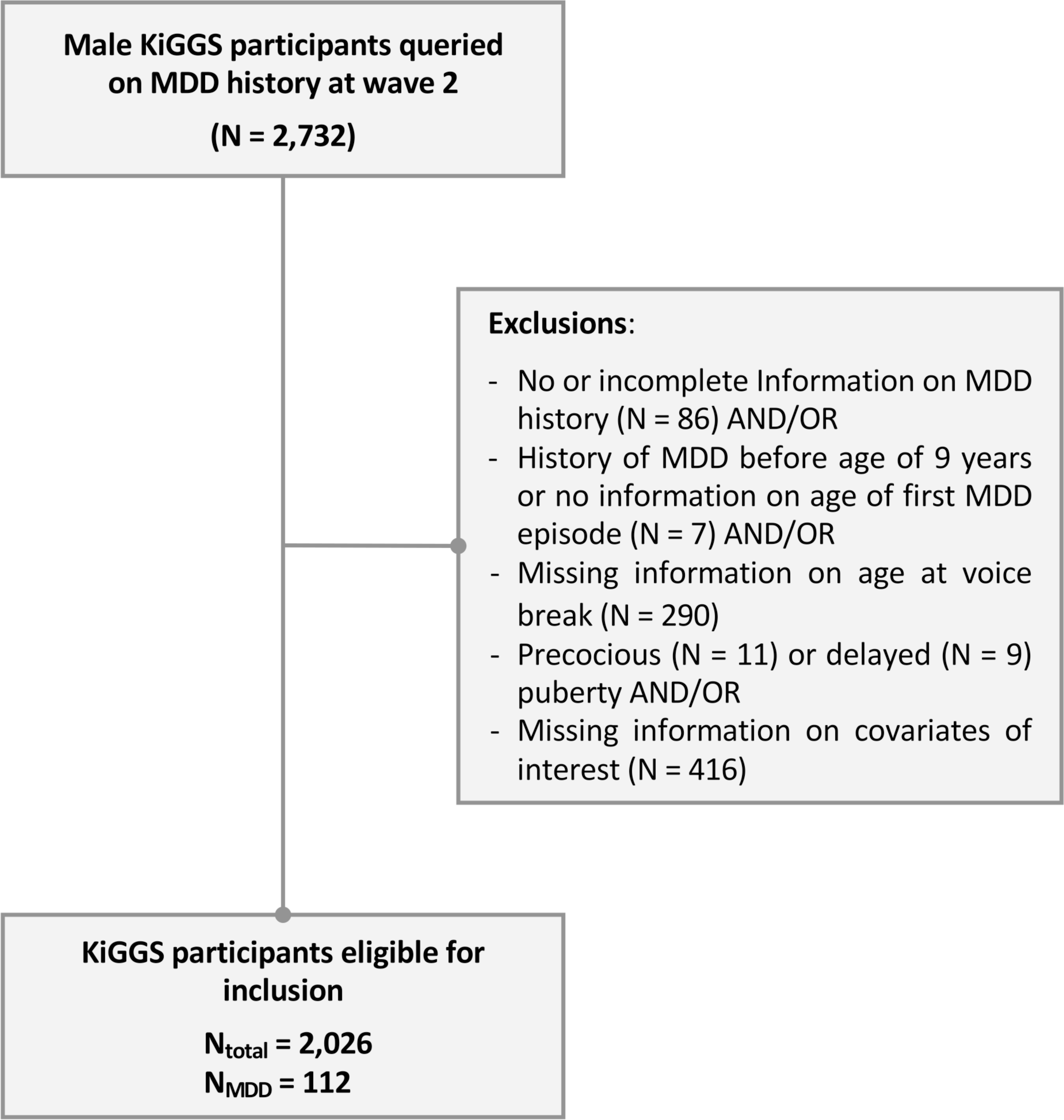
Study Design Flowchart. MDD = Major Depressive Disorder. Note: Some participants had missing information on more than more variable.

### Questionnaires and Interviews

Participants aged 11 years and older and parents of minors completed self-administered, standardized questionnaires. Among others, the questionnaires addressed physical and mental health as well as their determinants, including information on socioeconomic status (SES), social support, self-efficacy, and childhood trauma by standardized questionnaires (details on the questionnaires are provided in the Supplemental Methods 1). Moreover, participants were queried on their subjective body image (much too thin, slightly too thin, exactly the right weight, too obese, or much too obese) and on adverse childhood experiences (own severe disease or accident and prolonged hospital stay; severe disease or accident and death of a close person).

Parents of minors and young adults (≥ 18 years) additionally completed a computer-assisted personal interview (CAPI) conducted by a specially trained study physician (Mauz et al., 2020). Beginning with wave 2, the CAPI also covered the participants’ mental health history, including the age of onset of mental health conditions (in total years), regarding selected disorders diagnosed by a physician (general practitioner, psychiatrist, neurologist) or psychologist.

### Male Pubertal Timing and Physical Examination

MPT was defined by recalled age at voice break, considering its high correlation with other maturational pubertal events (Busch et al., 2019) and for consistency with the GWAS underlying the MR analysis. MPT was determined at wave 2 by retrospectively asking participants about the age at which their voice started to break (in total years) (Mauz et al., 2020). Based on this information, the total sample of male participants (N = 3,747) was used to define precious (mean age at voice break ≤ −2.5 SDS [0.4^th^ percentile]) and delayed puberty (≥ +2.5 SDS [99.6^th^ percentile]) (Pathomvanich, Merke, & Chrousos, 2000).

Further details regarding anthropometric measures in the psychiatric sample and KiGGS participants are outlined in the Supplementary Methods 2.

### Statistical Analysis – Significance, Effect Size, and Power

The results regarding the relationship between MPT and MDD were assessed by two-tailed testing and considered significant at p <.05. The results regarding the analysis of covariates, including those investigating interactions with MPT, were FDR-corrected for multiple comparisons at *q* <.05. The analysis of sociodemographic data and the pattern of missingness (see Supplementary Methods 3) were considered exploratory and not corrected for multiple testing.

The effect size of significant findings is reported as Cohen’s *d* (*d* [small 0.31 ≤ d ≤ 0.49, medium 0.50 ≤ *d* ≤ 0.79, large ≥ 0.8]). Post hoc power analyses were conducted with GPower 3.1.9.7 (Faul, Erdfelder, Lang, & Buchner, 2007), assuming a prevalence of MDD of 5%, α =.05, borderline small effect size (d = 0.2) per change in 1 SD of the predictor, and R^2^ of the predictor with other variables = 0.04.

### Logistic Regression Analysis

Three logistic regression models assessed the relationship between MPT (independent variable) and the risk for MDD (dependent variable). The first model was unadjusted, the second model accounted for information on confounder related to either MPT or MDD (BMI, SES, subjective body image, social support, self-efficacy, CTQ subscales emotional and physical abuse, and the sum score of childhood adverse events) recorded at wave 2, and the third model additionally considered interaction terms between subjective body image, social support, self-efficacy, CTQ subscales emotional and physical abuse, and childhood abuse and MPT (for details on testing the assumptions, see the Supplementary Methods 4).

## Mendelian Randomization

### Univariate MR Analysis

In a MR study, three main assumptions must be met (Haycock et al., 2016): (1) the genetic instrument must have a strong association with the exposure, (2) the genetic instrument is independent of potential confounders in the relationship between the exposure and the outcome, and (3) the outcome is associated with the genetic instrument only through the effect of the exposure (Haycock et al., 2016).

The first assumption can be tested by the F-statistic, calculated as (Beta/SE)^2^ for each SNP defining the IV for MPT. The F-statistic for the total IV including all SNPs was calculated considering the proportion of the explained variance by the IV (h^2^_SNP_) and the sample size of the outcome (Burgess, Small, & Thompson, 2017). An F > 10 indicates that no bias due to a weak IV is present (Haycock et al., 2016). The second assumption is unlikely to be violated in the MR context (Greco, Minelli, Sheehan, & Thompson, 2015), as genetic variants are fixed at conception and cannot be influenced by confounding factors of the risk factor-outcome associations (Haycock et al., 2016).

To test the validity of the third assumption, we used several approaches. First, we calculated Cochran’s Q-statistic to test for IV heterogeneity. In the univariate MR case, a significant finding (p < .05) indicates heterogeneity. Heterogeneity can have several causes, of which horizontal pleiotropy is the most likely (Greco et al., 2015). To specifically address directional unbalanced pleiotropy, we performed MR[Egger regression and MR-PRESSO (Mendelian Randomization Pleiotropy RESidual Sum and Outlier) analysis (Verbanck, Chen, Neale, & Do, 2018).

Considering that it is unlikely that all IVs fulfill the assumptions for instrumental variables, we followed recent recommendations (Burgess et al., 2019) and used several robust MR methods to account for violations of the MR assumptions (for details on the different methods, see the Supplementary Methods 5, available online). If Cochran’s Q-statistic suggested heterogeneity of the IV, the contamination mixture method and the MR-Lasso method were applied in addition to the penalized weighted median method.

For visualization of the results, Forest, scatter, funnel, and leave-one-out plots were created (see Supplementary Methods 5 and Figures 2-4).

**Figure 2.**
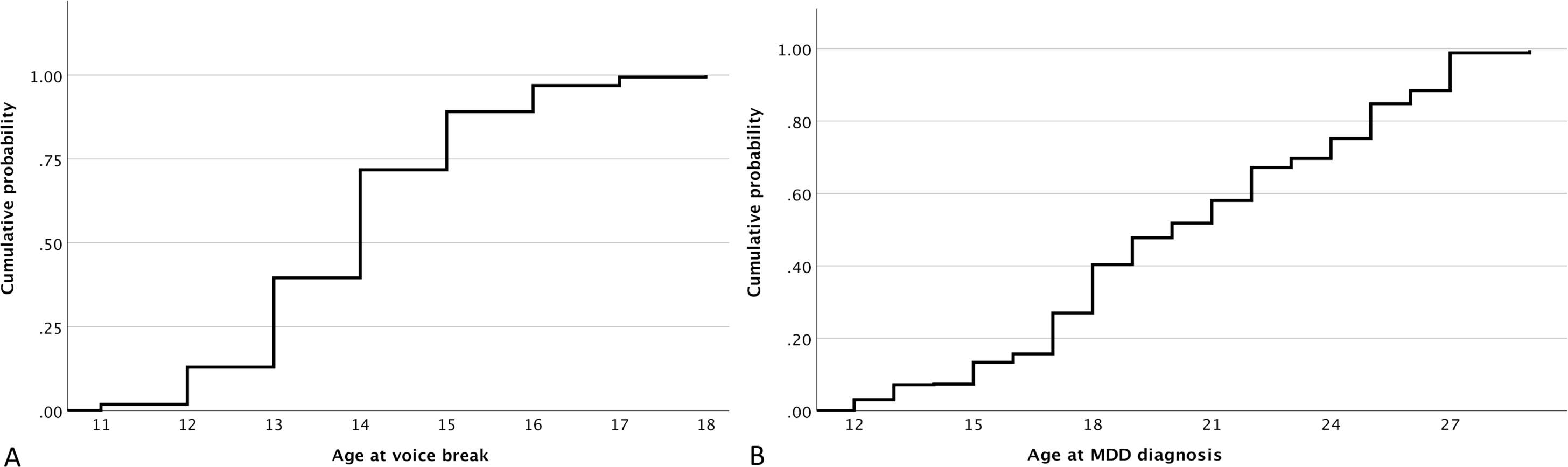
Cumulative probability of voice break (Panel A) and major depressive disorder (MDD, Panel B) dependent on age.

**Figure 3.**
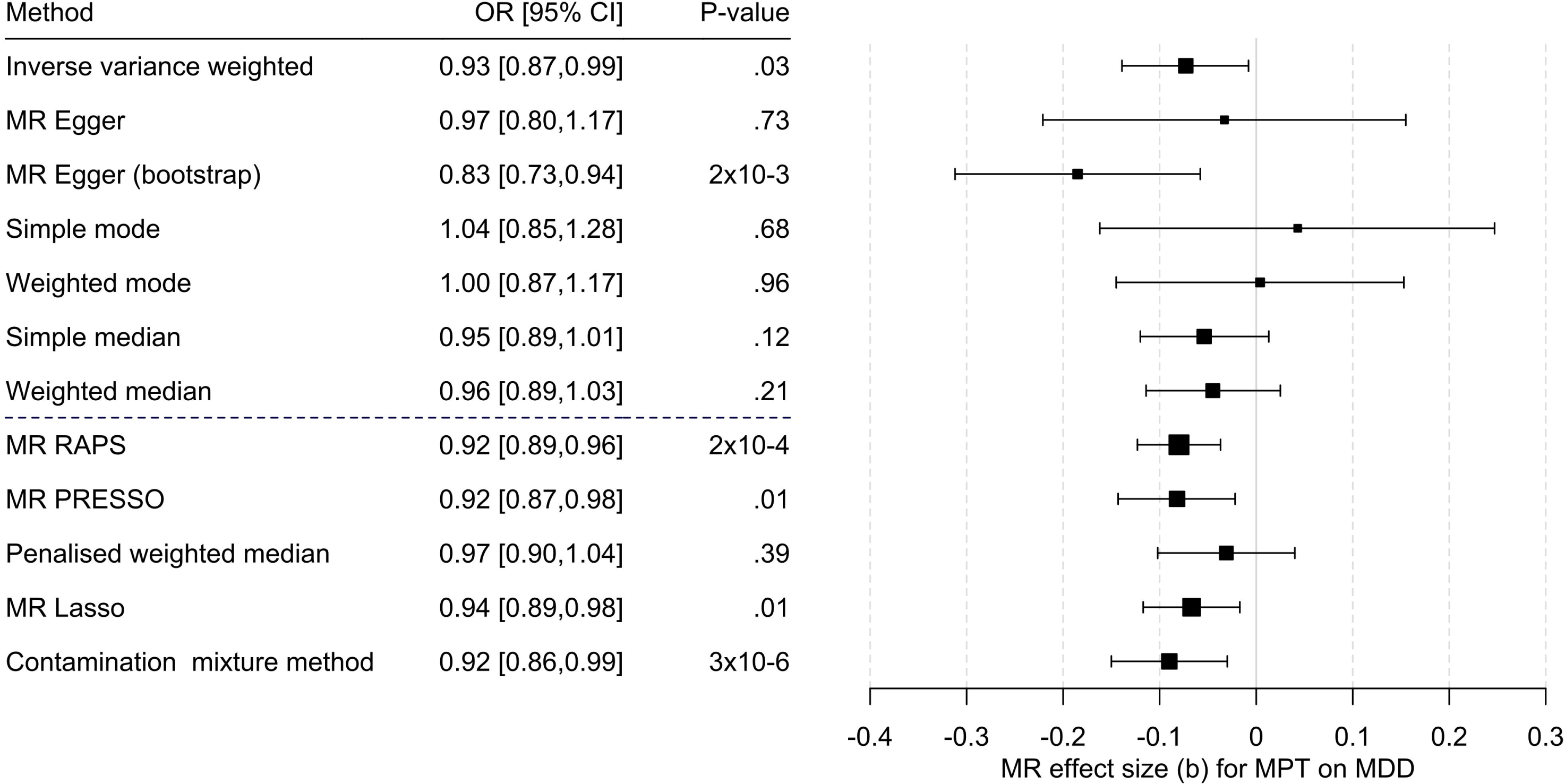
Results of the Multi-SNP Univariate Mendelian Randomization (MR) Analyses Regarding the Effect of Male Pubertal Timing (MPT) [Hollis et al. (2020)] on MDD Risk [Howard et al. (2019)]. OR = odds ratio, CI = confidence interval, b = unstandardized causal estimate of the change in risk for depression per one-year change in puberty time.

**Figure 4.**
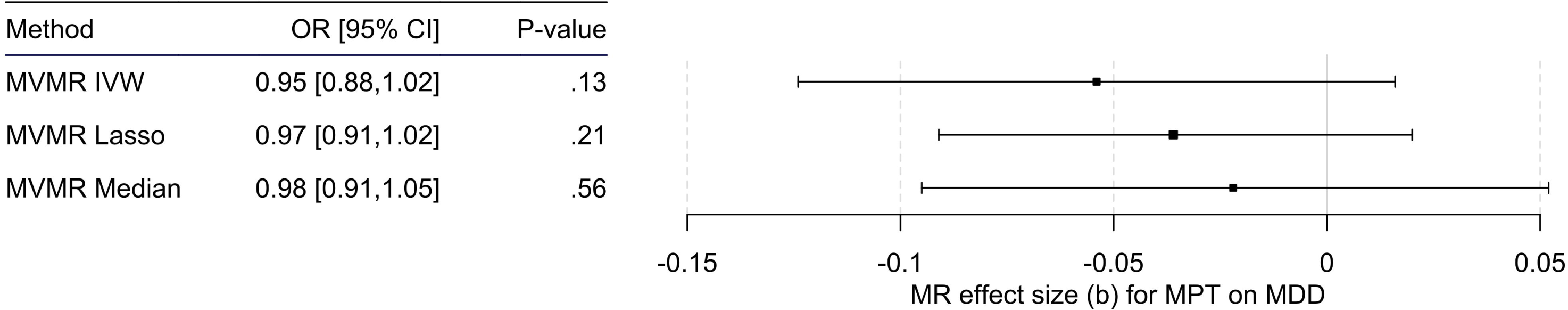
Results of the Multivariable MR (MVMR) Analyses of the Causal Effect of Male Pubertal Timing (MPT) [Hollis et al. (2020)] on MDD Risk [Howard et al. (2019)] Adjusted for BMI [Pulit et al. (2019)] Calculated Using Three Different Methods. OR = odds ratio, CI = confidence interval, b = unstandardized causal estimate of the change in risk for depression per one-year change in MPT.

Since SNP heritability was not reported by Hollis et al. (2020) for MPT, we estimated the explained variance by the 71 SNPs constituting our IV using the formula by Shim et al. (2015) for MR power analysis.

### Multivariable MR Analysis - Assessing the Effect of BMI and Other Causes of Pleiotropy

Previously, BMI has been identified as a potential pathway to horizontal pleiotropy when using pubertal timing as exposure, as a large number of SNPs associated with pubertal timing are also related to BMI (Day et al., 2017). This is supported by a recent MR study (Busch, Hojgaard, Hagen, & Teilmann, 2020), even though epidemiological evidence has not been consistent in this regard (Crocker et al., 2014; Lee et al., 2010). When also considering that a higher BMI is associated with a higher risk for depression (Casanova et al., 2021), BMI is a potential source of horizontal pleiotropy. Thus, to consider BMI as a confounder, we performed multivariable MR with four different methods (for details on the different methods, see the Supplementary Methods 5), including multivariable MR-PRESSO. If there was no evidence of pleiotropy by multivariable MR-PRESSO, we deemed it unlikely that further confounders needed consideration.

We further calculated the Q-statistic to test for heterogeneity. In contrast to the univariate case, a P value <.05 indicates that heterogeneity is unlikely in the multivariable MR model and that the selected SNPs can predict the exposure phenotypes (Sanderson, Spiller, & Bowden, 2020). As individual genetic data were not available, we could not consider correlations between the exposure variables.

### Male Pubertal Timing

To construct the instrument variable, we used the genome-wide significant independent SNPs (n=76) from the GWAS by Hollis et al. (2020). An effective GWAS meta-analysis sample size of 205,354 men of European ancestry was achieved by a multi-trait analysis of GWAS (MTAG) for continuous age at voice break by data obtained from the UK Biobank and 23andMe study (for further details on the GWAS, see the Supplementary Methods 6).

### Major Depressive Disorder

The data source for the outcome variable MDD was the recent GWAS by Howard et al. (2019) including 807,553 individuals (246,363 cases and 561,190 controls). This meta-analysis was based on data from the three largest genome-wide association studies of depression. These studies used different measures for depression: (a) self-reported clinical diagnosis of MDD by Hyde et al. (2016); (b) MDD obtained from a structured clinical interview or based on broader criteria by Wray et al. (2018); (c) self-reported help-seeking for problems with nerves, anxiety, tension or depression (broad depression) by Howard et al. (2019). Overlapping samples were excluded. The meta-analysis identified 102 independent genome-wide significant variants. The genome-wide SNP-based heritability (h^2^_SNP_) was 8.9%. The proportion of females was 48% in Hyde et al. (2016) and 54% in Howard et al. (2019). The proportion of males and females was not reported by Wray et al. (2018) (for further details on the GWAS, see the Supplementary Methods 6).

### BMI

We used the most recent GWAS on BMI with sex-specific analyses (Pulit et al., 2019). A total of 806,834 individuals of European ancestry were included in the analysis, 374.756 of whom were males. The SNP-based heritability (h^2^_SNP_) for males was 35% for all SNPs (Pulit et al., 2019).

### Reporting and Software

In reporting our studies, we followed the STROBE and STROBE-MR recommendations (Skrivankova et al., 2021). Data handling and analyses were either performed with SPSS 28.0 (IBM Corp., Armonk, NY) and its Complex Samples® procedures to account for the sample design, design-related effects, and attrition related to sociodemographic characteristics (Mauz et al., 2020) or the software ‘R’ (3.5.1 and 4.1.1) with the R-packages ‘TwoSampleMR’ (0.4.26, https://github.com/MRCIEU/TwoSampleMR) (Hemani et al., 2018), ‘Mendelian Randomization’ (0.5.1; https://CRAN.R-project.org/package=MendelianRandomization) (Broadbent et al., 2020), ‘MVMR’ (0.3; https://github.com/WSpiller/MVMR) (Sanderson et al., 2020), and ‘MRPracticals’ (0.0.1; https://github.com/WSpiller/MRPracticals). MR-PRESSO was performed with R-package (1.0; https://github.com/rondolab/MR-PRESSO) (Verbanck et al., 2018).

## Results

### Epidemiological Study - Results

Of the 17,640 children and adolescents who took part in the baseline assessment of the KiGGS study, 2,732 were male *and* were queried on MDD history at wave 2. Of these, 2,026 participants were considered for further analyses regarding the cross-sectional analysis, as 706 were excluded for reasons outlined in Figure 1.

### Missingness

Only regarding the history of MDD (unweighted: 3.1%, weighted 5.2%), MPT (weighted 13.7%, unweighted: 10.6%), and adverse life events (weighted 9.9%, unweighted: 9.0%) was there a significant (weighted) proportion (> 5%) of participants with missing information. Further analyses showed a random pattern of missingness.

Excluded participants were found to have a lower SES (p <.001), lower levels of social support (p =.004), lower self-efficacy (p =.02), and a history of more emotional childhood trauma (p =.04). However, absolute differences were minor (Table S1), and effect sizes were either borderline small (SES: d = 0.22) or negligible (d <.20).

### Descriptives

The mean age of eligible participants at wave 2 was 23.93 (SD 3.35) years, and the mean age at voice break was 13.9 (SD 4.5) years (Table 1, Figure 2A). The mean difference between the age at wave 2 and the age at voice break was 9.6 (SD 3.7) years. Considering a significant (p =.01) but trivial effect (d =.14) regarding the relationship between age at wave 2 and the reported age at voice break, no recall bias was evident.

**Table 1.**
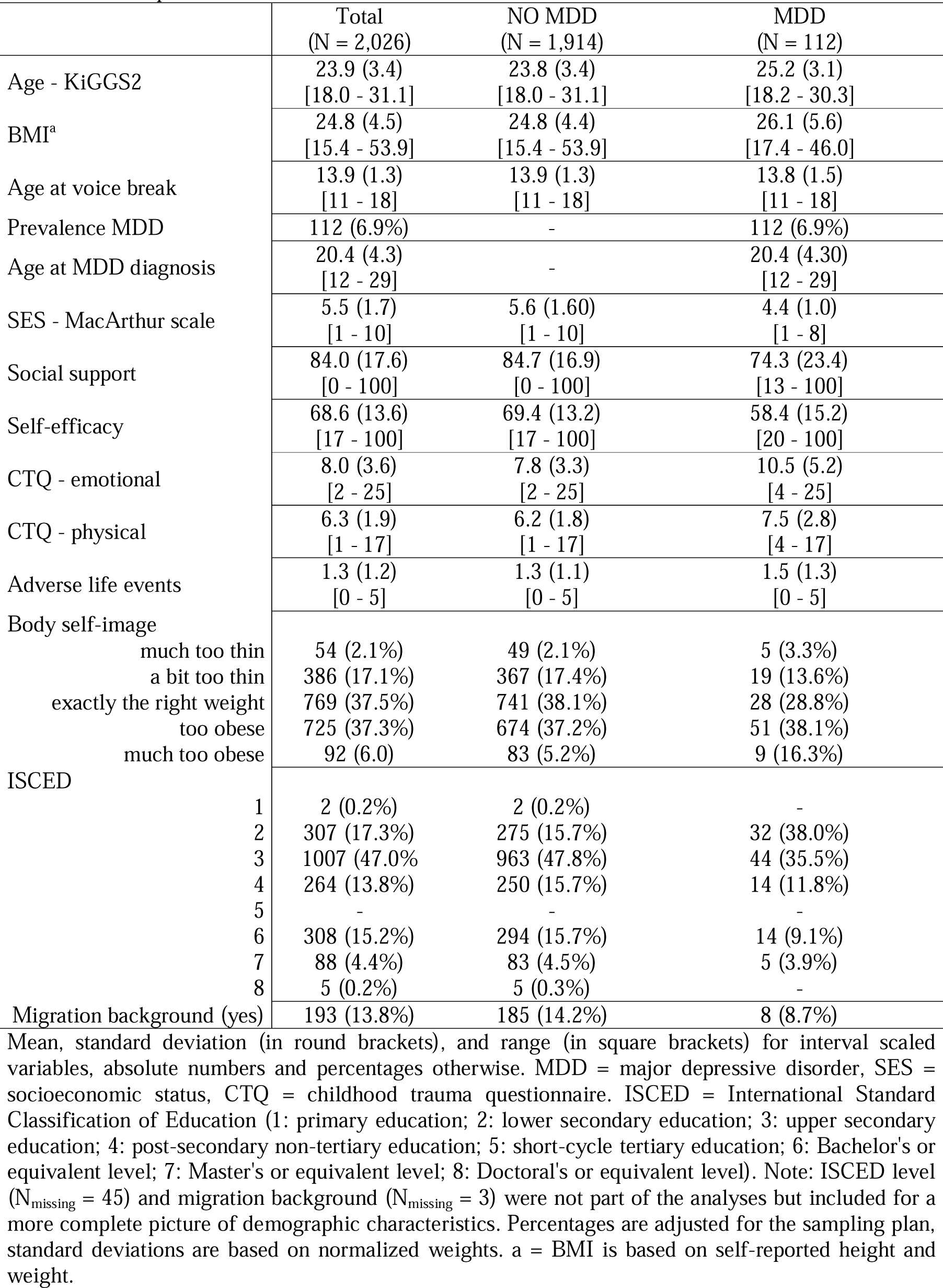
Descriptives

Over a median follow-up period of 11 years (IQR: 10-14), 112 cases of MDD were reported, of which only 25.9% were diagnosed before the age of 18 (Figure 2B).

### Logistic Regression Results

Neither in the unadjusted (b = −0.05, t = −0.46, OR = 0.95, 95%-CI [0.76 – 1.19], p = .65) nor the adjusted (b = 0.01, t = 0.06, OR = 1.01, 95%-CI [0.81 – 1.26], p = .93) model considering important confounders in the relationship between MPT and MDD risk was there a significant finding (Table S2 for detailed results). This did not change when additionally considering interactions between MPT and confounders (b = 1.12, t = 1.08, OR = 3.07, 95%-CI [0.40 – 23.74], p = .28). Power (1-β) to detect a borderline small effect size (d ≥ 0.2) was .95.

There was no evidence of a curvilinear relationship between MPT and MDD by the Box-Tidwell approach in either model.

### Mendelian Randomization - Results

For the univariable MR on the effect of MPT on MDD risk, 71 SNPs could be considered for the IV (Table S3. The F-statistic for the IV was 16685.2. The F-statistic for each SNP individually indicated that all SNPs were sufficiently strong instruments (lowest F value = 29.9, Table S4).

Cochran’s Q test indicated that the IV was heterogenic (MR-Egger – Q(df = 69) = 186.4, p = 9.5 x 10^-13^; IVW - Q(df = 70) = 187.0, p = 1.3 x 10^-12^). Consistently, MR-PRESSO indicated pleiotropy even though Egger’s intercept did not (= −0.001, SE = 0.003, p =.66), likely because MR-PRESSO is more sensitive to horizontal pleiotropy than Egger’s intercept (Verbanck et al., 2018).

Univariable MR showed mixed results: IVW and MR-Egger with bootstrapping showed a significant causal effect of MBT on MDD. The methods robust to pleiotropy (MR RAPS, outlier corrected MR-PRESSO) and two of three methods robust to heterogeneity (contamination mixture method, MR Lasso) also showed a significant effect (Figure 3, Table S5, Figures S2-S4). However, the multivariable MR of the effect of PBT on risk for MDD (n = 71; Table S6), adjusted for BMI, showed that the effect observed in the univariable MR could be due to pleiotropy related to BMI (Table S7 Figure 4), as none of the three multivariable methods showed an effect of MPT on MDD risk. Multivariate MR-PRESSO did not identify pleiotropic outliers. The conditional F-test showed that the IV was sufficiently strong for both MPT (F_TS_ = 39.3) and BMI (F_TS_ = 27.0). The Q-statistic confirmed the validity of the IV (Q(df = 68) = 180.6; p = 3.7 x 10^-12^), as there was no evidence of significant heterogeneity.

### Power Analysis

The h^2^ for the 71 SNPs associated with MPT was 2.1% (Table S4). Our analysis had a power of 80% to detect an OR of 0.95 or 1.05 for MDD per one-year change in MPT and 100% power to detect an OR of 0.92 or 1.08 (Figure S1).

## Discussion

Previous studies have provided inconsistent findings regarding the association of MPT with depressive symptoms and MDD (Hamlat et al., 2014), likely for methodological reasons. However, when investigating this relationship with a large epidemiological study considering multiple potential confounders and within the MR framework robust to residual confounding and reverse causation to consider the methodological limitations of each approach, there was no evidence of a (causal) relationship in this regard in the present work.

### Male Pubertal Timing and MDD

The relationship between MPT and depression has mainly been studied dimensionally, that is, regarding the severity of depressive symptoms on a continuous scale. These studies have provided mixed evidence (Hamlat et al., 2014), reporting a relationship between depressive symptoms and either earlier or later pubertal timing, off-timing effects (i.e., effects of earlier and later pubertal timing), or no relationship. These inconsistencies also apply to those few studies addressing this relationship categorically, that is, the presence (or absence) of MDD according to clinical criteria, as investigated in the present study. While confounders were not considered in either study, Angold et al. (1998) found no evidence of a relationship between pubertal timing in girls and boys, but Hamlat et al. (2020) identified pubertal timing as a risk factor for incident and recurrent MDD in both sexes. However, considering the present study’s findings, the latter study was likely affected by residual confounding. Moreover, this study may have also been insufficiently powered to conclusively investigate the impact of pubertal timing on MDD risk in boys, as discussed by the authors. In contrast, the present epidemiological analyses and the MR study were sufficiently powered for sound conclusions. However, for valid conclusions of the MR, the genetic architecture of depression should not change throughout the life course. As previously discussed in more detail (Hirtz et al., 2022), a recent genome-wide association meta-analysis showed that the genetic architecture of internalizing symptoms is stable from early childhood to adolescence. This was concluded from the observation of overlapping SNP-based heritability estimates during earlier and later life and a high genetic correlation between childhood internalizing symptoms and adult depression (*r_g_* > 0.7) (Jami et al., 2022).

### Closely Related but Yet Distinct – Pubertal Timing in Girls and Boys

The genetic SNP-based underpinnings of pubertal timing in females and males largely overlap (Hollis et al., 2020). Of the 76 SNPs related to MPT, 71 show a similar effect estimate of the same direction observed for age at menarche. This could imply that pubertal timing in both sexes share the same underlying mechanisms to implicate sexual maturation in mental health. In females, downstream mechanisms to age at menarche have been suggested to explain its effect on MDD (Magnus et al., 2020). Discussed mechanisms include, for example, younger age at first delivery, an increased risk of sexual abuse, and younger age at first sexual intercourse (Magnus et al., 2020). In particular, the latter is consistent with risky sexual behavior in adolescent boys with a small to medium effect size in a recent meta-analysis (Ullsperger & Nikolas, 2017) and a significant genetic correlation between MPT and health risk behaviors, including alcohol consumption and smoking (Hollis et al., 2020). However, at the same time, early MPT is associated with favorable social traits, including educational attainment (Hollis et al., 2020), which does not seem to apply to girls (Torvik et al., 2021). Moreover, physical changes related to pubertal maturation in boys are often considered desirable and convey social value, in contrast to such changes in adolescent girls (Rudolph & Troop-Gordon, 2010). Thus, adverse outcomes regarding mental health conferred by the genotype underlying pubertal timing might be counterbalanced by advantageous effects related to the same set of genes and their individual allelic architecture in males but not in females. This would explain the findings of the present study as well as the observation of adverse effects of earlier pubertal timing in girls on the mental health phenotype (Hirtz et al., 2022).

However, the present study also provides insights into the underlying mechanisms relating pubertal timing to the etiology of MDD in boys. From a genetic perspective, BMI drives the apparent relationship between MPT and MDD due to an overlapping genetic origin of both exposure phenotypes. This is indicated by the multivariable MR analysis and the observation of no cause of horizontal pleiotropy other than via BMI. In this regard, BMI has not only been related to pubertal timing by epidemiological and MR studies (Busch et al., 2020) but has also been implicated in MDD (Casanova et al., 2021). BMI may exert its effects on MDD risk via several downstream outcomes, including, for example, hormone levels (Milano et al., 2020), peer relations (Kanders, Nilsson, & Åslund, 2021), and body image (Richard, Rohrmann, Lohse, & Eichholzer, 2016). In contrast, in the epidemiological analyses in the present study, neither BMI nor body image were related to MDD risk. However, since MR studies assess the average effect of an exposure over the life course, the direct and indirect effects of BMI may no longer be apparent or have yet to manifest in an epidemiological context.

### Limitations – Epidemiological Study

The age at voice break was self-reported in the epidemiological (and genetic) study. However, there was no evidence of a substantial recall bias. Moreover, self-assessed age at voice break has been shown to correlate closely with other important pubertal events, justifying its use in epidemiological studies (Busch et al., 2019).

In neither the epidemiological analyses nor the MR study, we could consider subclinical depressive symptoms. As a consequence, our conclusions are limited to overt depression. Moreover, confounders other than those included in the epidemiological analyses have been suggested but were not captured at either KiGGS assessment. However, issues of residual confounding are sufficiently covered by the MR analyses, confirming the results of the epidemiological studies. Also, neither migration nor social status is related to MPT in the KiGGS sample, rendering them unlikely confounders (Kahl, Rosario, & Schlaud, 2007).

### Limitations – MR study

By the statistical model of the GWAS by Hollis et al. (2020) underlying the MR analysis, we were limited to studying a linear relationship between MPT and MDD. However, there was no evidence of a relationship other than linear by the epidemiological study.

The analysis of the effect of BMI on MPT in the multivariable MR model relied on the GWAS by Pulit et al. (2019). While this GWAS allowed us to study sex-specific BMI-related findings pertaining only to males, it was conducted in adults. This implies a continuity of the genetic architecture of BMI over the life course regarding the present study, which is supported by a recent meta-analysis of 26 GWASs on childhood BMI. This study included data from 61,111 children and identified 25 childhood-specific BMI loci (Vogelezang et al., 2020). Most of these SNPs (20 of 25) were also related to adult BMI, and there was a genetic correlation between childhood and adult BMI of *r_g_* = 0.76.

The genetic instrument for depression was not sex specific, as no such information is available. However, as previously discussed (Hirtz et al., 2022), a recent study that evaluated between-sex genetic heterogeneity in MDD using GWAS summary statistics from 29 cohorts found a genetic correlation close to one (Trzaskowski et al., 2019).

As previously mentioned, the covariance between AAM and BMI could not be considered in the multivariable MR analysis because individual data were not available. Moreover, our results apply to the Caucasian population only and likely do not generalize to patients with precocious puberty, even though SNPs in genes related to precocious puberty were used as IV as well (Hollis et al., 2020).

## Conclusions and Implications

In contrast to females, our study shows that earlier MPT is not related to an increased MDD risk. This is suggested by the epidemiological and genetic analyses, combining methodological advantages of both approaches, which allows for sound conclusions. Instead of pubertal timing, the MR study demonstrates that BMI is a driving force increasing MDD risk. It remains to be determined at what age this effect is most influential. However, considering the multitude of adverse health outcomes related to higher BMI levels, the findings of our study support the rationale for preventive measures at the individual and population level to address obesity and its related risk factors. Our findings also suggest that practitioners should focus on BMI and its peripubertal changes rather than on the timing of puberty when assessing the risk profile for MDD in adolescent boys.

## Data availability

The reported results related to the KiGGS study are based on a secondary analysis of data provided by the Robert Koch Institute (RKI), Germany. Requests to access the datasets should, therefore, be directed to the RKI.

Publicly available datasets analyzed in this study can be found here: https://www.ebi.ac.uk/gwas/. Original data generated and analyzed during this study (i.e., harmonized genetic data) are part of the online Supplementary Material.

## Author contribution statement

RH and TP: Conceptualization, Methodology, Formal analysis, Writing. CG: Conceptualization and Writing – Review & Editing. HH: Investigation (KiGGS) and Writing – Review & Editing. NA, AH, JH: Methodology and Writing – Review & Editing. All authors participated in critically revision of the manuscript for important intellectual content. All authors approved the final version to be published.

## Conflicts of interest

The authors report no financial or other relationship relevant to the subject of this article.

## Data Availability

The reported results related to the KiGGS study are based on a secondary analysis of data provided by the Robert Koch Institute (RKI), Germany. Requests to access the datasets should, therefore, be directed to the RKI. Publicly available datasets analyzed in this study can be found here: https://www.ebi.ac.uk/gwas/. Original data generated and analyzed during this study (i.e., harmonized genetic data) are part of the online Supplementary Material.

https://doi.org/10.6084/m9.figshare.21856791.v1

## Acknowledgements

The authors also acknowledge support from the Open Access Publication Fund of the University of Duisburg-Essen. The funders had no role in the study design, data collection and analysis, decision to publish, or manuscript preparation.

## Ethical standards

The authors assert that all procedures contributing to this work comply with the ethical standards of the relevant national and institutional committees on human experimentation and with the Helsinki Declaration of 1975, as revised in 2008.

## Notes

### Competing Interest Statement

The authors have declared no competing interest.

### Funding Statement

This study did not receive any funding.

### Author Declarations

The ethics committee of the Charite Berlin (baseline and wave 1) and the ethics committee of the Hannover Medical School (wave 2) gave ethical approval of the KiGGS study. Details on the ethical approval concerning the GWAS used are provided in the respective study.

